# Risk and severity of COVID-19 and ABO blood group in transcatheter aortic valve patients

**DOI:** 10.1101/2020.06.13.20130211

**Authors:** Marion Kibler, Adrien Carmona, Benjamin Marchandot, Kensuke Matsushita, Antonin Trimaille, Mohamad Kanso, Laurent Dietrich, Cécile How-Choong, Albane Odier, Gabrielle Gennesseaux, Ophélie Schramm, Antje Reydel, Michel Kindo, Minh Hoang, Sébastien Hess, Chisato Sato, Sophie Caillard, Laurence Jesel, Olivier Morel, Patrick Ohlmann

**Affiliations:** Division of Cardiovascular Medicine, Strasbourg University Hospital, Strasbourg, France; INSERM (French National Institute of Health and Medical Research), UMR 1260, Regenerative Nanomedicine, FMTS, Strasbourg, France; Division of Nephrology and Transplantation, Strasbourg University Hospital, Strasbourg, France; Division of Cardiac Surgery, Strasbourg University Hospital, Strasbourg, France

**Author notes:** **Correspondence:** Patrick Ohlmann, MD, PhD, Pôle d’Activité Médico-Chirurgicale Cardiovasculaire, Nouvel Hôpital Civil, BP 426 - 67091, Strasbourg, France.

**Keywords:** ABO blood group, Coronavirus Disease 2019, Transcatheter Aortic Valve Replacement

## Abstract

**Background:** Although cardiovascular disease has been associated with an increased risk of coronavirus disease 2019 (COVID-19), no studies have reported its clinical course in patients with aortic stenosis who had undergone transcatheter aortic valve replacement (TAVR).

Several observational studies have found an association between the A blood group and an increased susceptibility to SARS-CoV-2 infection, whereas the O blood group appears to be protective.

**Objective:** To investigate the frequency and clinical course of COVID-19 in a large sample of patients who had undergone TAVR and to determine the associations of the ABO blood group with disease occurrence and outcomes.

**Methods:** Patients who had undergone TAVR between 2010 and 2019 were included in this study and followed-up through the recent COVID-19 outbreak. The main outcomes were the occurrence and severity (hospitalization and/or death) of COVID-19 and their association with the ABO blood group.

**Results:** Of the 1125 patients who had undergone TAVR, 403 (36%) died before January 1, 2020, and 20 (1.8%) were lost to follow-up. The study sample therefore consisted of 702 patients. Among them, we identified 22 cases (3.1%) with COVID-19. Fourteen patients (63.6%) were hospitalized or died of disease. Multivariate analysis identified the A blood group (*versus* others) as the only independent predictor of COVID-19 in patients who had undergone TAVR (odds ratio [OR] = 6.32; 95% confidence interval [CI] = 2.11-18.92; p=0.001). The A blood group (*versus* others; OR = 8.27; 95% CI = 1.83-37.43, p=0.006) and a history of cancer (OR = 4.99; 95% CI = 1.64-15.27, p = 0.005) were significantly and independently associated with disease severity (hospitalization and/or death).

**Conclusions:** Patients who had undergone TAVR are vulnerable to COVID-19. The subgroup with the A blood group was especially prone to develop the disease and showed unfavorable outcomes.

**Condensed abstract:** Among 702 patients who had undergone TAVR between 2010 and 2019 and who were alive on January 1, 2020, 22 patients developed COVID-19. Fourteen patients (63.6%) were hospitalized or died of disease. The A blood group (*versus* others) was the only independent predictor of COVID-19. The A blood group and a history of cancer were significantly and independently associated with disease severity (hospitalization and/or death). Altogether these findings suggest that patients who had undergone TAVR are vulnerable to COVID-19. The subgroup with the A blood group was especially prone to develop the disease and showed unfavorable outcomes.

## INTRODUCTION

At the end of 2019, a new zoonotic coronavirus (SARS-CoV-2) responsible of coronavirus disease 2019 (COVID-19), arose from Wuhan, Hubei Province, China. The World Health Organization (WHO) on March 11, 2020, has declared COVID-19 outbreak a global pandemic. In fact, SARS-CoV-2 spread rapidly in 166 other countries around the world, resulting in a global burden of 4 170 424 laboratory-confirmed cases and a death toll of 287 399 as of May 14, 2020.^(1)^. The Alsace region in eastern France has been significantly impacted, resulting in fast reshaping of in-hospital facilities. Several cardiology divisions have been converted into dedicated COVID-19 units – with cardiac care units being repurposed as intensive care units (ICUs) ^(2,3)^.

A history of cardiovascular (CV) disease is currently recognized as a risk factor for the occurrence and severity of COVID-19, especially in the elderly ^(4,5)^. Accordingly, previous studies have indicated that up to 40% of patients who require ICU admission for COVID-19 had preexisting congestive heart failure; further, the mortality rate from COVID-19 for patients with preexisting CV disease may be as high as 36% ^(6)^. While there is ample literature to suggest a direct role for a history of heart disease in the susceptibility and severity of COVID-19, its clinical course in patients with valvular disease remains poorly investigated. With a growing number of patients with aortic stenosis being treated with transcatheter aortic valve replacement (TAVR), there is a strong need to investigate this interaction further. Much of the recent focus in COVID-19 research has revolved around biological markers of disease susceptibility and/or severity. The ABO blood group has been shown to affect individual vulnerability to SARS-CoV ^(7)^, hepatitis B virus ^(8)^, Norwalk virus ^(9)^, and *Helicobacter pylori* infection ^(10)^. Notably, observational studies have found an association between the A blood group and an increased susceptibility to SARS-CoV-2 infection while the O blood group appears to be protective ^(11–13)^.

The present study was undertaken to evaluate the clinical course of COVID-19 in patients with aortic stenosis who had undergone TAVR. We also examined whether the ABO blood group is associated with susceptibility to and severity of COVID-19 in this clinical population, and whether this association is independent of potential confounders.

## METHODS

### Study setting and patient enrollment

This study consists of a retrospective, observational investigation aimed at examining the occurrence and severity of COVID-19 in a large population of patients who undergone TAVR for severe aortic stenosis between 2010 and 2019. The study was conducted in the Strasbourg University Hospital (Strasbourg, Alsace, eastern France). The general characteristics of the study patients – including demographics, medical history, echocardiography findings, and ABO blood group – were determined from their medical records and entered into an electronic file along with follow-up data. During the COVID-19 outbreak, all patients were contacted by phone to ascertain their health status, cardiovascular and COVID-19 symptoms, medication use, and outcomes. Patient-reported data collected through a standardized questionnaire were thoroughly cross-checked with official clinical records. The study was reviewed and approved by the Institutional Review Board at the Strasbourg University Hospital (CE-2020-69). Owing to the retrospective nature of the study, the need for informed consent was waived.

### Definitions

In accordance with WHO technical guidance ^(14)^, patients were considered as confirmed cases of COVID-19 in presence of positive reverse transcriptase-polymerase chain reaction (RT-PCR) testing of a nasopharyngeal swab specimen. Because RT-PCR can yield false-negative results, patients with typical symptoms and characteristic imaging findings on chest computed tomography (CT) were classified as confirmed cases ^(15)^. Patients who were hospitalized for or died of COVID-19 were considered to have severe disease.

### Statistical analysis

Descriptive statistics are expressed as means ± standard deviations for continuous data or as counts (percentages) for categorical variables. Survival curves according to the ABO blood group were plotted by the Kaplan-Meier method (log-rank test) with right censoring at the time of last follow-up (May 8, 2020). The time-to-event was calculated as the time elapsed from January 1, 2020, to the date of the index event (disease onset, hospitalization, or death). Logistic regression models were constructed to evaluate the unadjusted and covariate-adjusted odds ratios (ORs) and 95% confidence intervals (CIs) for the occurrence of COVID-19, COVID-19-related death, and severe COVID-19. Variables adjusted for in the multivariate models were those showing univariate associations at a p value <0.20. Statistical analyses were performed using SPSS, version 17.0 (IBM, Armonk, NY, USA). All tests were two-sided, and statistical significance was set as a p value of <0.05.

## RESULTS

### General characteristics

Between 2010 and 2019, a total of 1125 patients with aortic stenosis underwent TAVR in our hospital. We excluded 423 patients from the analysis due to death before January 1, 2020 (n = 403) or loss to follow-up (n = 20). Figure 1 depicts the flow of participants through the study. The general patient characteristics (n = 720; mean age: 82 ± 6.9 years; 44% men) are provided in Table 1. Common coexisting CV comorbidities included coronary artery disease (45.3%), atrial fibrillation (40.3%), congestive heart failure (35.9%), and peripheral arterial disease (27.2%). A positive history of cancer was present in 26.9% of cases, whereas chronic obstructive pulmonary disease and chronic kidney disease were identified in 11.7% and 16.5% of the study patients, respectively. At the time of interview, CV medications included angiotensin converting enzyme (ACE) inhibitors/angiotensin II receptor blockers (48.9%), statins (50.2%), anticoagulants (45.4%), and aspirin (53.3%).

**Table 1.** General characteristics of patients who had undergone transcatheter aortic valve replacement according to the presence or absence of COVID-19

| **Clinical Characteristics** | **Entire cohort**<br>**(n= 702)** | **COVID-19**<br>**(n= 22)** | **No COVID-**<br>**19 (n= 680)** | **p**<br>**value** |
| --- | --- | --- | --- | --- |
| Age, years | 82 ± 6.9 | 82 ± 8.4 | 82 ± 6.9 | 0.961 |
| Male sex – n (%) | 313 (44) | 7 (31.8) | 306 (45) | 0.220 |
| STS score – % | 5.9 ± 4.9 | 5.5 ± 2.4 | 5.9 ± 5.0 | 0.757 |
| **Cardiovascular risk factors – n (%)** |  |  |  |  |
| Current smoking | 26 (3.7) | 1 (4.5) | 25 (3.7) | 0.832 |
| Hypertension | 587 (83.6) | 18 (81.8) | 569 (83.7) | 0.817 |
| Obesity (Body mass index > 30 kg/m<sup>2</sup>) | 183 (26.1) | 6 (27.3) | 177 (26.1) | 0.899 |
| Dyslipidemia | 428 (61) | 12 (54.5) | 416 (61.2) | 0.530 |
| Diabetes | 213 (30.3) | 6 (27.3) | 207 (30.4) | 0.750 |
| **Comorbidities– n (%)** |  |  |  |  |
| Coronary artery disease | 318 (45.3) | 12 (54.5) | 306 (45.0) | 0.376 |
| Congestive heart failure | 252 (35.9) | 6 (27.3) | 246 (36.5) | 0.392 |
| Stroke | 98 (14) | 3 (13.6) | 95 (14.0) | 0.964 |
| Atrial fibrillation | 283 (40.3) | 6 (27.3) | 277 (40.7) | 0.205 |
| Peripheral arterial disease | 191 (27.2) | 5 (22.7) | 186 (27.4) | 0.631 |
| COPD | 82 (11.7) | 3 (13.6) | 79 (11.6) | 0.740 |
| Prior cancer | 189 (26.9) | 10 (45.5) | 179 (26.3) | 0.053 |
| CKD (Creatinine levels >130 µmol/l) | 115 (16.5) | 4 (18.2) | 111 (16.4) | 0.824 |
| LVEF after TAVR– % | 56 ± 11 | 56 ± 12 | 56 ± 11 | 0.902 |
| **Treatment at time of follow up – n (%)** |  |  |  |  |
| Aspirin | 365 (53.3) | 13 (59.1) | 352 (53.1) | 0.579 |
| **P2Y12 inhibitors** |  |  |  |  |
| VKA | 144 (21.0) | 4 (18.2) | 140 (21.1) | 0.740 |
| DOAC | 175 (25.5) | 6 (27.3) | 169 (25.5) | 0.850 |
| ACE-i/ARB | 335 (48.9) | 12 (54.5) | 323 (48.7) | 0.591 |
| Statins | 344 (50.2) | 9 (40.9) | 335 (50.5) | 0.375 |
| Amiodarone | 101 (14.7) | 2 (9.1) | 99 (14.9) | 0.447 |
| **ABO blood type – n (%)** |  |  |  |  |
| A | 299 (42.6) | 18 (81.8) | 281 (41.3) | 0.002 |
| B | 63 (9) | 0 (0) | 63 (9.3) |  |
| AB | 20 (2.9) | 0 (0) | 20 (2.9) |  |
| O | 320 (45.6) | 4 (18.2) | 316 (46.5) |  |
| Rhesus positive (Rh+) – n (%) | 352 (58.6) | 11 (68.8) | 341 (58.3) | 0.402 |
| **Blood type – no. (%)** |  |  |  |  |
| A Rh- | 87 (12.4) | 3 (13.6) | 84 (12.4) | 0.027 |
| A Rh+ | 212 (30.2) | 15 (68.2) | 197 (29.0) |  |
| AB Rh- | 8 (1.1) | 0 (0) | 8 (1.2) |  |
| AB Rh+ | 17 (2.4) | 0 (0) | 17 (2.5) |  |
| B Rh- | 14 (2.0) | 0 (0) | 14 (2.1) |  |
| B Rh+ | 49 (7.0) | 0 (0) | 49 (7.2) |  |
| O Rh- | 74 (10.5) | 1 (4.5) | 73 (10.7) |  |
| O Rh+ | 154 (21.9) | 2 (9.1) | 152 (22.4) |  |
| Missing | 87 (12.4) | 1 (4.5) | 86 (12.6) |  |

Data are given as means ± standard deviations or counts (percentages). Abbreviations: ACE-i: Angiotensin Converting Enzyme inhibitor; ARB: Angiotensin Receptor blocker; CKD: Chronic Kidney Disease (creatinine >
130 µmol/l); COPD: Chronic Obstructive Pulmonary Disease; COVID-19: Coronavirus Disease 2019; DOAC: direct oral anticoagulant; LVEF: Left Ventricular Ejection Fraction; STS score: Society of Thoracic Surgeons score; TAVR: Transcatheter Aortic Valve Replacement; VKA: vitamin K antagonist.

**Figure 1.**
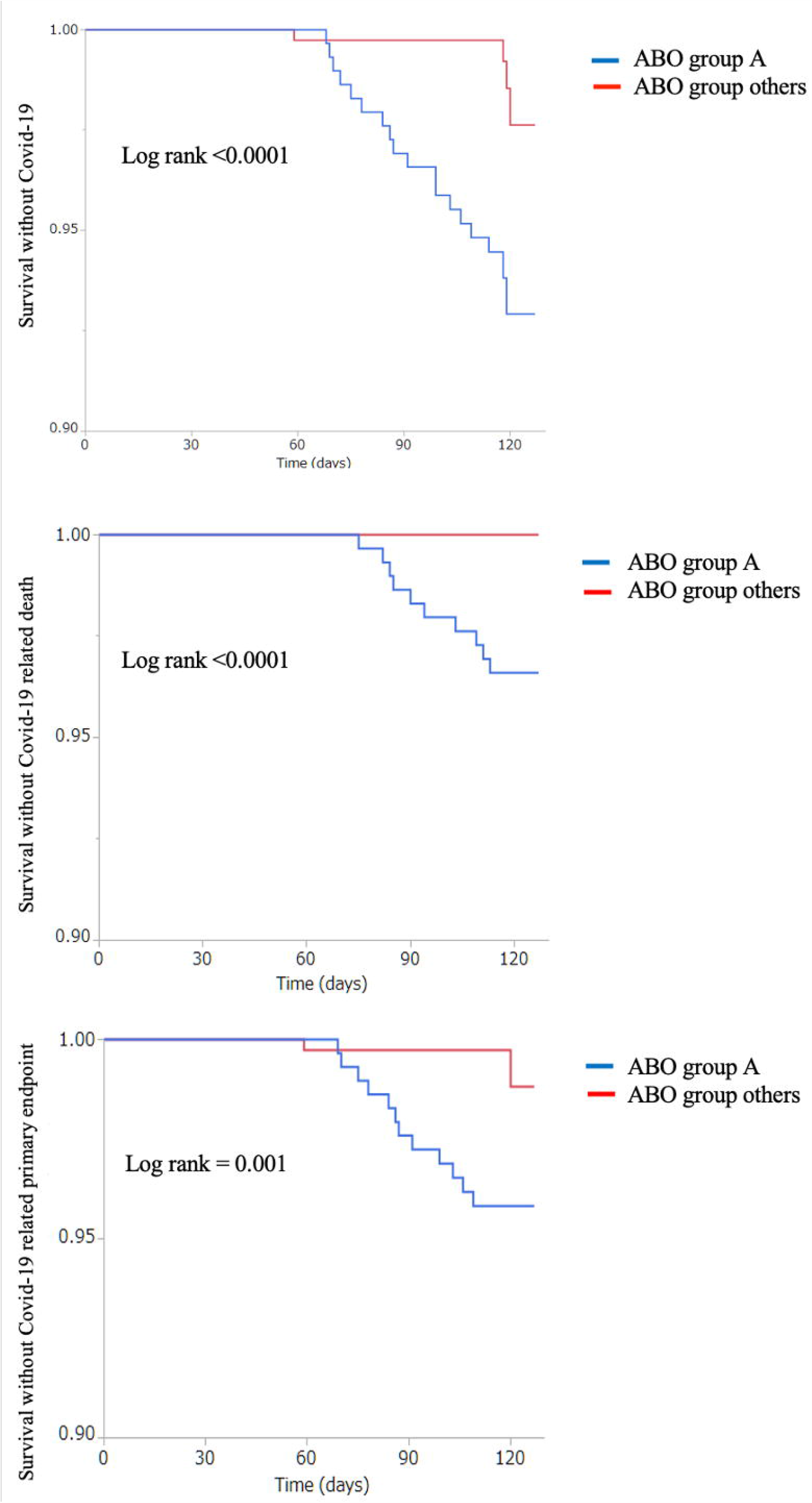
Flow of patients through the study. Abbreviations: CT: Computed tomography, Covid-19: Coronavirus Disease 2019; ICU: Intensive Care Unit; RT-PCR: Reverse-Transcriptase-Polymerase-Chain-Reaction; TAVR: Transcatheter Aortic Valve Replacement.

### Occurrence and presentation of COVID-19

Eighty-two patients (11.4%) had suspected COVID-19. Of them, 61 underwent RT-PCR testing and 21 chest CT. Diagnosis was confirmed in 22 cases (3.1%; 21 by RT-PCR and one on chest CT). Of them, 14 (63.6%) were hospitalized or died of COVID-19. Common clinical symptoms at presentation included dyspnea (77.3%), fever (77.3%), and cough (72.7%). Myalgia, gastrointestinal manifestations, and anosmia/ageusia occurred in 40.9%, 27.3%, and 18.2% of participants, respectively.

### COVID-19, hospitalizations, and mortality

As of January 1, 2020, the all-cause and cardiovascular mortality rates in the study patients were 6.8% and 2.8%, respectively. Compared with patients without COVID-19, those with the disease had significantly higher all-cause mortality (5.6% *versus* 45.5%, respectively; p <0.001) and hospitalization (1.8% *versus* 59.1%, respectively; p <0.0001) rates (Table 2).

**Table 2.** Clinical outcomes of patients who had undergone transcatheter aortic valve replacement according to the presence or absence of COVID-19

|  | **Entire cohort**<br>**(n = 702)** | **COVID-19**<br>**(n = 22)** | **No COVID-19 (n = 680)** | **p value** |
| --- | --- | --- | --- | --- |
| **Hospitalization – n (%)** | 25 (3.6) | 13 (59.1) | 12 (1.8) | <0.0001 |
| **Conventional unit** | 22 (3.2) | 10 (45.5) | 12 (1.8) | <0.0001 |
| **Intensive care unit** | 3 (0.44) | 3 (13.6) | 0 (0) | <0.0001 |
| **Mortality from January 1, 2020 – n (%)** |  |  |  |  |
| **All-cause mortality** | 48 (6.8) | 10 (45.5) | 38 (5.6) | <0.0001 |
| **Cardiovascular mortality** | 20 (2.8) | 0 (0) | 20 (2.8) | 0.414 |
| **COVID-19 mortality** | 10 (1.5) | 10 (45.5) | 0 (0) | <0.0001 |
| **COVID-19 severity – n (%)** |  |  |  |  |
| **COVID-19 related hospitalization or death** | 14 (2.0) | 14 (63.6) | 0 (0) | <0.0001 |

Abbreviations: COVID-19: Coronavirus Disease 2019.

### COVID-19 and ABO blood group

Patients with COVID-19 more frequently had the A blood group than those without (81.8% *versus* 41.3%, respectively). Conversely, the O (18.2% *versus* 46.5%, respectively), B (0% *versus* 9.3%, respectively), and AB (0% *versus* 2.9%, respectively) groups were underrepresented in patients with COVID-19. Subgroup analyses were subsequently performed according to the Rhesus (Rh) group. Interestingly, the A Rh+ blood type (68.2% *versus* 29%, respectively) – but not the A Rh-type (13.6 *versus* 12.4%, respectively) – was overrepresented in patients with COVID-19. The O Rh+ (9.1% *versus* 22.4%, respectively), O Rh-(4.5% *versus* 10.7%, respectively), B Rh+ (0% *versus* 7.2%, respectively), B Rh- (0% *versus* 2.1%, respectively), AB Rh+ (0% *versus* 2.5%, respectively), and AB Rh- (0% *versus* 1.1%, respectively) types were all underrepresented in patients with COVID-19 (Table 1). Additional analyses were also performed according to blood group A. Patients with the A blood group were more likely to develop COVID-19 compared to those with other blood types (6 *versus* 1%, respectively; p < 0.0001). In addition, with respect to others blood groups, patients with the A blood group presented more frequently COVID-19-related death (3.4 *versus* 0%, respectively; p < 0.0001) as well as the combined endpoint of COVID-19-related death or hospitalization (4 *versus* 0.5%, respectively; p < 0.001; Supplemental Table 1).

### Predictors of COVID-19

A history of cancer and the blood type A were significant predictors of COVID-19 in univariate analysis. Multivariate analysis identified the A blood group (*versus* others) as the only independent predictor of COVID-19 in patients who had undergone TAVR (OR = 6.32; 95% CI = 2.11−18.92; p=0.001; Table 3). Kaplan-Meier plots of COVID-19-free survival according to the blood group (A *versus* others) are shown in Figure 2, panel A.

**Table 3.** Factors associated with the occurrence of COVID-19 in patients who had undergone transcatheter aortic valve replacement

|  | Univariate analysis |  |  | Multivariate analysis |  |  |
| --- | --- | --- | --- | --- | --- | --- |
|  | OR | 95% CI | p value | OR | 95% CI | p value |
| Age | 0.99 | 0.94-1.06 | 0.610 |  |  |  |
| Male sex | 0.57 | 0.23-1.42 | 0.226 |  |  |  |
| Diabetes | 0.86 | 0.33-2.22 | 0.751 |  |  |  |
| Obesity | 0.89 | 0.41-2.76 | 0.899 |  |  |  |
| Hypertension | 0.89 | 0.29-2.64 | 0.817 |  |  |  |
| Dyslipidemia | 0.76 | 0.32-1.79 | 0.531 |  |  |  |
| Current smoking | 0.83 | 0.16-9.65 | 0.832 |  |  |  |
| Atrial fibrillation | 0.55 | 0.21-1.41 | 0.212 |  |  |  |
| Peripheral artery disease | 0.78 | 0.28-2.15 | 0.632 |  |  |  |
| CKD (Creatinine levels >130 µmol/l) | 1.13 | 0.38-3.41 | 0.703 |  |  |  |
| Prior cancer | 2.33 | 0.99-5.49 | 0.053 | 2.28 | 0.96-5.43 | 0.062 |
| ACE-i/ARBs | 1.26 | 0.54-2.96 | 0.591 |  |  |  |
| P2Y12 inhibitors | 0.70 | 0.09-5.37 | 0.736 |  |  |  |
| Aspirin | 1.28 | 0.54-3.03 | 0.580 |  |  |  |
| Statins | 0.68 | 0.29-1.61 | 0.377 |  |  |  |
| A blood group | 6.29 | 2.14-19.08 | 0.001 | 6.32 | 2.11-18.92 | 0.001 |

Abbreviations: ACEi, angiotensin converting enzyme inhibitors; ARB, angiotensin receptor blockers; CI, confidence interval; CKD, chronic kidney disease; COVID-19, coronavirus disease 2019; LVEF, left ventricular ejection fraction; OR, odds ratio; TAVR, transcatheter aortic valve replacement.

**Figure 2.**
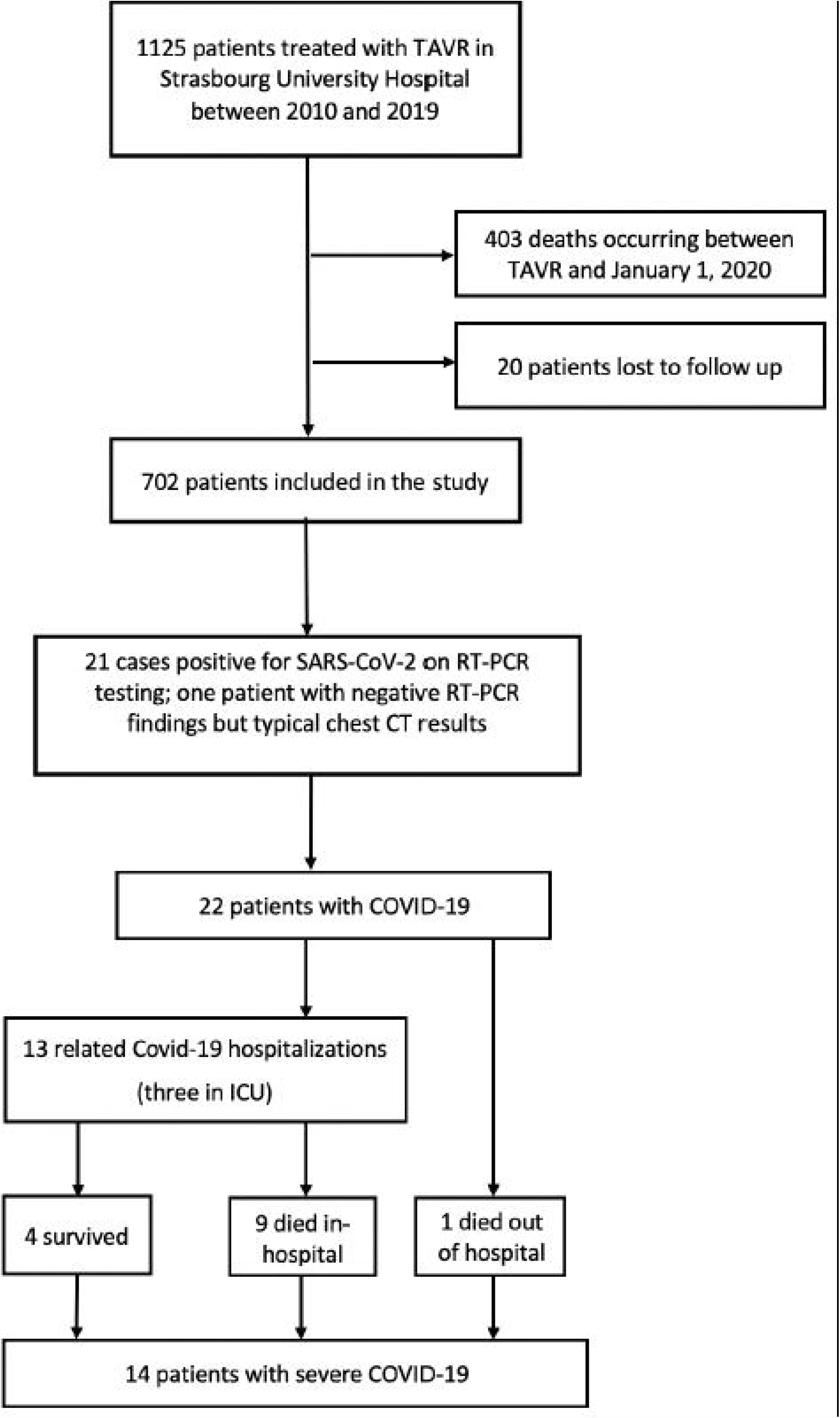
Kaplan–Meier plots of COVID-19-free survival (panel A), COVID-19-related mortality (panel B), and severe-COVID-19-free survival (panel C) according to the ABO blood group (group A *versus* other groups).

### Predictors of severe COVID-19

Multivariate analysis (Table 4) revealed that the blood group A (*versus* others; OR = 8.27; 95% CI = 1.83−37.43, p=0.006) and a history of cancer (OR = 4.99; 95% CI = 1.64−15.27, p = 0.005) were significantly and independently associated with COVID-19 severity (hospitalization and/or death). Kaplan-Meier plots of COVID-19-related mortality and severe-COVID-19-free survival are shown in Figure 2 (panels B and C, respectively).

**Table 4.** Factors associated with severe COVID-19 in patients who had undergone transcatheter aortic valve replacement

|  | Univariate analysis |  |  | Multivariate analysis |  |  |
| --- | --- | --- | --- | --- | --- | --- |
|  | OR | 95% CI | p value | OR | 95% CI | p value |
| Age | 0.98 | 0.92-1.05 | 0.649 |  |  |  |
| Sex (male) | 0.68 | 0.23-2.07 | 0.502 |  |  |  |
| Diabetes | 1.74 | 0.59-5.09 | 0.309 |  |  |  |
| Obesity | 0.77 | 0.21-2.78 | 0.688 |  |  |  |
| Hypertension | 0.71 | 0.19-2.59 | 0.608 |  |  |  |
| Dyslipidemia | 0.85 | 0.29-2.48 | 0.767 |  |  |  |
| Atrial fibrillation | 0.57 | 0.18-1.69 | 0.371 |  |  |  |
| Peripheral artery disease | 1.07 | 0.33-3.47 | 0.908 |  |  |  |
| CKD (Cr >130 µmol/l) | 2.07 | 0.64-6.71 | 0.226 |  |  |  |
| Coronary artery disease | 1.63 | 0.56-4.74 | 0.373 |  |  |  |
| Heart failure | 0.71 | 0.22-2.29 | 0.566 |  |  |  |
| COPD | 1.26 | 0.28-5.75 | 0.761 |  |  |  |
| Stroke | 1.03 | 0.23-4.66 | 0.972 |  |  |  |
| Prior cancer | 5.08 | 1.68-15.34 | 0.004 | 4.99 | 1.64-15.27 | 0.005 |
| A blood group | 8.38 | 1.86-37.74 | 0.006 | 8.27 | 1.83-37.43 | 0.006 |
| O blood group | 0.19 | 0.04-0.87 | 0.033 |  |  |  |
| Aspirin | 0.87 | 0.30-2.52 | 0.804 |  |  |  |
| ACE-i/ARB | 1.05 | 0.36-3.01 | 0.934 |  |  |  |
| Statins | 0.74 | 0.25-2.15 | 0.579 |  |  |  |

Abbreviations: ACEi, angiotensin converting enzyme inhibitors; ARB, angiotensin receptor blockers; CI, confidence interval; CKD, chronic kidney disease; COPD, chronic obstructive pulmonary disease; COVID-19, coronavirus disease 2019; DOAC, direct oral anticoagulants; LVEF, left ventricular ejection fraction; OR, odds ratio; STS, Society of Thoracic Surgeons; TAVR, transcatheter aortic valve replacement; VKA, vitamin K antagonists.

## DISCUSSION

To our knowledge, this study is the first to specifically investigate the impact of COVID-19 in patients who have undergone TAVR. There are two principal findings from our research. First, patients who had undergone TAVR were at high risk to contract COVID-19. Second, the A blood group was identified as a significant risk factor for both the occurrence and severity of COVID-19.

### Prevalence of COVID-19

The prevalence of COVID-19 in our patients who had undergone TAVR was 3.13%, which is higher than that observed in the general French population (24/10 000 on May 14, 2020) ^(16)^. Whether the increased COVID-19 rate was due to local characteristics of the outbreak in Alsace or to a higher susceptibility conferred by prior CV disease ^(17)^ needs further epidemiological study. Moreover, the mortality rate from COVID-19 was 45% in the current investigation. Age, frailty, and a significant burden of comorbidities are possible explanations for the high death toll ^(18)^. Moreover, 63.6% of our patients had severe disease (hospitalization and/or death). Despite these findings, the mechanisms by which CV disease confers susceptibility to COVID-19 remain unclear. Evidence is emerging on the association between aggressive disease and loss of ACE-2 function as a result of its proteolytic cleavage ^(19)^.

Under physiological conditions, ACE-2 counteracts the detrimental effects of angiotensin II ^(20)^, which might be overexpressed in patients with CV disease ^(21)^. At sites of endothelial injury, an imbalance between ACE-2 and ACE-1 activity may result in angiotensin II accumulation – which further exacerbates tissue injury and promotes both inflammation and thrombosis ^(22)^. Although our study does not address the role of ACE-2 in the susceptibility to COVID-19 among patients who had undergone TAVR, its involvement is certainly plausible.

### ABO blood group and COVID-19

In the current study, patients who had undergone TAVR and had the A blood group were more prone to develop COVID-19 and were more likely to experience unfavorable outcomes. In a research conducted in 2173 Chinese patients, Zhao et al. ^(23)^ showed for the first time that the A blood group was associated with an increased disease susceptibility to COVID-19 while the O group seemed less vulnerable. They also found higher death rates in patients with the A group. A more recent report from the Central Hospital of Wuhan confirmed the increased risk conferred by the A group and the reduced disease susceptibility associated with the O group ^(12)^. Another study connecting the A blood group with an increased risk of contracting COVID-19 is a research on 1599 individuals who underwent SARS-CoV-2 testing in the United States ^(24)^. However, no relation with in-hospital mortality was found. The authors carried these observations a step further with Rh antigen testing and found that its expression could modulate the association of the ABO blood group with disease susceptibility. Similar findings were noticed in our study, albeit limited to the A group. If a subject had the A group and was also Rh+, the patient would have a substantially higher risk of COVID-19, whereas this was not the case for Rh-individuals.

Growing evidence indicates that the A blood group is associated with an increased susceptibility to and severity of COVID-19. The mechanisms beyond this association are unknown but several hypotheses might be raised. It is possible that anti-A antibodies could lead to a decreased interaction of SARS-CoV-2 with its cellular receptor ACE-2 ^(25)^. Interestingly, the A blood group has also been related with an increased risk of CV disease ^(26)^. Numerous biological pathways have been proposed to account for the association between the A blood group and atherothrombosis – including an increased production of soluble intercellular adhesion molecules ^(27)^ and/or Von Willebrand factor (vWF) ^(28)^. Other authors have emphasized the significance of vWF cleavage in subjects with the O blood group ^(29)^ – an event which may reduce thrombotic risk in SARS-CoV-2-infected individuals ^(13)^.

### Limitations

Several caveats of our investigation need to be considered. First, our study employed a retrospective design and the number of observed events (deaths and/or hospitalizations) was limited. As such, the presence of residual confounding may pose limitations in our ability to generalize our conclusions. Second, our research has an exploratory nature and we cannot rule out the presence of chance findings resulting from multiple comparisons. Another caveat is that the sex distribution of participants varied across the ABO blood groups. Specifically, men were underrepresented in the A blood group, potentially posing limitations in our ability to fully explore the impact of this variable on COVID-19 severity.

## Conclusion

Patients who had undergone TAVR are vulnerable to COVID-19. The subgroup with the A blood group was especially prone to develop the disease and showed unfavorable outcomes. Our results add to the growing literature indicating that the ABO blood group may be a useful laboratory parameter that should be taken into account for risk stratification during clinical work-up of patients with COVID-19.

### Clinical perspectives

#### Competency in patient care and procedural skills

patients who had undergone TAVR are vulnerable to COVID-19. The subgroup with the A blood group was especially prone to develop the disease and showed unfavorable outcomes.

### Translational outlook

particular attention should be payed to patients who have undergone TAVR, to prevent COVID-19 and anticipate the severity of the disease. ABO group may be an additional tool for risk stratification in these patients.

Patients who have undergone TAVR are vulnerable to Covid-19

## Data Availability

The database is available online with the link below

https://www.dropbox.com/s/cdny37mtajzp0cs/BDD%20COVID%20TAVI%20V1.xlsx?dl=0

## Acknowledgements

This work was made possible by the front-line healthcare personnel of the Department of Cardiology, Strasbourg University Hospital. Their dedication during this unprecedented health crisis has been invaluable.

## Disclosure of potential conflicts of interest

The authors have no conflicts of interest in relation to the findings reported in this article.

## Sources of funding and support

This study was financially supported by GERCA (Groupe pour 237 l’Enseignement, la prévention et la Recherche Cardiologique en Alsace).

## Abbreviations

ACE-2: Angiotensin converting enzyme 2
COVID-19: coronavirus disease 2019
CT: Computed tomography
CV: Cardiovascular
Rh: Rhesus
RT-PCR: Reverse transcriptase Polymerase Chain reaction
SARS-CoV-2: Severe Acute Respiratory Coronavirus-2
TAVR: Transcatheter aortic valve replacement
WHO: World Health Organization

---

**Supplemental Table 1.** Clinical outcomes of patients who had undergone transcatheter aortic valve replacement according to A blood group

| Variable | Entire cohort<br>(n = 702) | A blood group<br>(n = 299) | Others blood groups (n = 403) | p value |
| --- | --- | --- | --- | --- |
| **Confirmed COVID-19** | 22 (3.1) | 18 (6) | 4 (1) | <0.0001 |
| **Mortality** |  |  |  |  |
| COVID-19 mortality | 10 (1.5) | 10 (3.4) | 0 (0) | <0.0001 |
| **COVID-19 severity – n (%)** |  |  |  |  |
| COVID-19 related hospitalization or death | 14 (2.0) | 12 (4.0) | 2 (0.5) | 0.001 |

Abbreviations: Covid-19: Coronavirus Disease 2019.

**Supplemental Table 2.** General characteristics of patients who had undergone transcatheter aortic valve replacement according to the presence or A blood group versus other groups.

| Variable | Entire cohort<br>(n = 702) | A blood group<br>(n = 299) | Other blood groups<br>(n = 403) | p value |
| --- | --- | --- | --- | --- |
| Age, years | 82.6 ± 6.9 | 82.4 ± 7.1 | 82.8 ± 6.9 | 0.513 |
| Male sex – n (%) | 313 (44.6) | 116 (38.8) | 197 (48.9) | 0.008 |
| STS score – % | 5.9 ± 5.0 | 6.0 ± 5.1 | 5.8 ± 4.9 | 0.645 |
| Cardiovascular risk factors – n (%) |  |  |  |  |
| Current smoking | 26 (3.7) | 8 (2.7) | 18 (4.5) | 0.214 |
| Hypertension | 587 (83.6) | 253 (84.6) | 334 (82.9) | 0.539 |
| BMI (kg/m<sup>2</sup>) | 27.3 ± 5.8 | 27.3 ± 5.8 | 27.2 ± 5.6 | 0.892 |
| Dyslipidemia | 428 (61.0) | 186 (62.2) | 242 (56.5) | 0.562 |
| Diabetes | 213 (30.3) | 97 (32.4) | 116 (28.8) | 0.297 |
| Comorbidities – n (%) |  |  |  |  |
| Coronary artery disease | 318 (45.3) | 146 (48.8) | 172 (42.7) | 0.106 |
| Congestive heart failure | 252 (35.9) | 107 (35.8) | 145 (36.0) | 0.958 |
| Stroke | 98 (14.0) | 40 (13.4) | 58 (14.4) | 0.701 |
| Atrial fibrillation | 283 (40.3) | 121 (40.5) | 162 (40.2) | 0.943 |
| COPD | 82 (11.7) | 36 (12.1) | 46 (11.4) | 0.786 |
| Prior cancer | 187 (26.6) | 82 (27.4) | 105 (26.1) | 0.685 |
| CKD (Creatinine levels >130 µmol/l) | 102 (14.6) | 50 (16.8) | 52 (12.9) | 0.150 |
| Peripheral arterial disease | 191 (27.2) | 84 (28.1) | 107 (26.6) | 0.650 |
| Blood Type – n (%) |  |  |  |  |
| A | 299 (42.6) | 299 (100) | 0 (0) | < 0.0001 |
| B | 63 (9.0) | 0 (0) | 63 (15.6) |  |
| AB | 20 (2.8) | 0 (0) | 20 (5.0) |  |
| O | 320 (45.6) | 0 (0) | 320 (79.4) |  |
| Treatment at the time of follow up – n (%) |  |  |  |  |
| Aspirin | 365 (53.3) | 163 (54.9) | 202 (52.1) | 0.463 |
| P2Y12 inhibitors | 44 (6.3) | 19 (6.4) | 25 (6.2) | 0.935 |
| VKA | 144 (21.0) | 58 (19.5) | 86 (22.2) | 0.401 |
| DOAC | 175 (25.5) | 78 (26.3) | 97 (25.0) | 0.707 |
| ACE-i/ARB | 335 (48.9) | 152 (51.2) | 183 (47.2) | 0.298 |
| Statins | 344 (50.2) | 156 (52.5) | 188 (48.5) | 0.291 |

Data are given as means ± standard deviations or counts (percentages). Abbreviations: ACEi, angiotensin converting enzyme inhibitors; ARB, angiotensin receptor blockers; BMI: Body mass index; CKD, chronic kidney disease; COPD, chronic obstructive pulmonary disease; COVID-19, coronavirus disease 2019; DOAC, direct oral anticoagulants; LVEF, left ventricular ejection fraction; STS, Society of Thoracic Surgeons; TAVR, transcatheter aortic valve replacement; VKA, vitamin K antagonists.

## Notes

### Competing Interest Statement

The authors have declared no competing interest.

### Funding Statement

No external funding

### Author Declarations

The study was reviewed and approved by the Institutional Review Board at the Strasbourg University Hospital (CE-2020-69)

